# A Latent Variable Approach for the Investigation of the Multi-factor Impact on Sleep Problems in Individuals with Mild Cognitive Impairment

**DOI:** 10.1101/2025.02.13.25322199

**Authors:** Chrysanthi Nega, Kleio Moustaka, Ion N. Beratis

## Abstract

The present study aims to explore the impact of neuropsychiatric symptoms, cognitive functioning, and medical status on sleep problems in individuals with Mild Cognitive Impairment (MCI), focusing on insomnia, by implementing a latent variable approach. The 131 Greek patients with MCI, who are consecutive visitors of the Alzheimer’s day clinic, underwent comprehensive assessments in cognitive functioning, neuropsychiatric symptoms, sleeping disturbances, and medical health status. The model was estimated by applying a Partial Least Squares (PLS) approach to structural equation modeling (SEM), incorporating the following latent variables: “psychological burden” (3 indicators: depressive symptoms, anxiety, and stress), “cognitive status” (5 indicators: general cognitive status, phonemic and semantic fluencies, psychomotor speed, and episodic memory), and “sleep problems” (5 indicators: overnight and earlier final awakening(s), decreased daytime functioning and well-being, and insufficient sleep duration). The results supported the SEM effectiveness, with good model fit and high predictive power. The “psychological burden” played the most significant role in the development of sleep problems within the clinical group of MCI, while “cognitive status” appeared to be the second most important SEM predictor, showing a negative association with “sleep problems”. Additionally, MCI age of onset and sleep problems severity were negatively associated. This latent-variable approach allowed us to capture the multidimensional complexity of sleep problems in MCI. Thus, applying integrative evaluations and interventions that target the cognitive status and psychological well-being, may improve the sleep quality of individuals with MCI, and further contribute to a slower cognitive decline course.

## Introduction

Mild Cognitive impairment (MCI) exists on the cognitive continuum between normal age-related decline and early-stage dementia. MCI does not define a future course of dementia, but it does indicate an increased risk compared to the general population to develop dementia of the Alzheimer’s type. It is characterized by cognitive deficits that do not significantly interfere with the ability to perform activities of daily living (ADLs). Subjective cognitive complaints (SCCs), such as feeling forgetful or experiencing a sense of mental decline, may be present but are not necessary or sufficient for diagnosis. The clinical detection of MCI relies on objectively identified deficits on cognitive testing adjusted for the individual’s age and education. According to Petersen et al. (20029), these deficits can manifest as amnestic MCI (aMCI), characterized by memory impairment entailing a higher risk of conversion to AD, or non-amnestic MCI (naMCI), involving cognitive difficulties in areas other than memory. Typically, deficits are limited to one (single-domain MCI; sdMCI) or a few cognitive domains (multi-domain MCI; mdMCI), such as executive function/processing speed, language, or learning and memory. Differential diagnosis between MCI and mild dementia primarily focuses on the preserved ability to perform ADLs in MCI. While some individuals with MCI may experience mild difficulty with complex activities, particularly work-related tasks, their overall daily function remains largely intact, and autonomy is preserved (Cornelis et al., 2017; Lu et al., 2023).

Sleep problems are a major neuropsychiatric symptom frequently observed in individuals with MCI. The prevalence of sleep problems is significantly higher in people with MCI compared to cognitively healthy older adults, with studies showing up to 70% of individuals with MCI being affected (Beaulieu-Bonneau & Hudon, 2009; da Silva, 2015; Song et al., 2020). In addition, older adults with MCI exhibit different patterns in sleep problems compared with normal elderly, with the former group experiencing shorter sleep duration and lower habitual sleep efficiency (An et al., 2014). A systematic review has also shown that significant changes in the frequency and severity of sleep problems mirror the progression of cognitive impairment, among healthy older adults, and older adults with MCI and AD (Casagrande et al., 2022). Growing evidence links sleep problems, including insomnia characterized by difficulty falling asleep, night awakenings, early morning awakenings, and non-restorative sleep (Cross et al, 2019), to a greater risk of MCI and dementia (Xu et al., 2020). These problems, particularly insomnia, play a significant role in the progression of cognitive decline (Um et al., 2022). Insomnia is associated with a 27% higher risk of cognitive disorders, including MCI (Xu et al., 2020), and individuals with insomnia are more prone to exhibit significant impairments in attention and episodic memory (Hung et al. 2018). Furthermore, both short and long sleep durations, as well as sleep quality issues (e.g., fragmentation, prolonged latency, excessive time in bed, apnea), contribute to cognitive impairment, including MCI, showing an inverted U-shape relationship between daily sleep duration and all-cause cognitive decline (Franks et al., 2021; Xu et al., 2020). Concerning the clinical subtypes of MCI, research is inconclusive on whether individuals with aMCI experiencing sleep problems are at a higher risk of developing dementia compared to those with naMCI and sleep problems. Even though previous studies have focused mostly on aMCI, evidence has shown no differences between aMCI and naMCI in sleep problems, as these were measured by Pittsburgh Sleep Quality Index (PSQI) scores (McKinnon et al., 2014). Still, other research indicated an association between sleep problems and naMCI, suggesting a potential subtype-specific relation between sleep problems and subjective sleep quality in the aforementioned clinical group (Rozzini et al., 2017). Given that aMCI is more strongly linked to the development of Alzheimer’s disease (Levey et al., 2006), whereas non-amnestic subtypes may progress to a variety of disease outcomes (e.g., vascular dementia, Lewy body dementia) (Petersen et al., 2001), understanding the subtype-specific relationships between sleep problems and MCI is crucial to understand disease trajectory and implement appropriate interventions.

Depression and anxiety are also common neuropsychiatric symptoms in individuals with MCI, often worsening sleep problems, which can further contribute to cognitive decline. These factors, along with hypertension, obesity, smoking, and lack of physical exercise are known risk factors for cognitive decline (Norton et al., 2014). The prevalence of clinically significant depression in individuals with MCI ranges from 20% in community-dwelling people to 83% in hospitalized samples (Gallagher et al., 2017; Ismail et al., 2017; Martin & Velayudhan, 2020), while the prevalence rates of anxiety range from 11.6% in population-based samples to 26.3% in clinical samples (Chen et al, 2018; Martin & Velayudhan, 2020). Research investigating the association between depression and anxiety and sleep problems has yielded mixed results. Yu et al. 2017 in a sample of participants with MCI in Asia, found that poor sleep quality directly contributes to cognitive decline, independently of depression and anxiety symptoms. Furthermore, DiNapoli et al. (2017) found that, while self-reported sleep quality did not correlate with objective sleep measures, participants with negative sleep discrepancy (worse self-reported sleep than actigraphy readings) had significantly higher depression scores. Similarly, Rozzini et al. (2017), classified individuals with MCI into good and bad sleepers and found that those classified as bad sleepers exhibited a significantly higher frequency of subjective depressive and anxious symptoms. Additionally, even depressed mood is strongly correlated with sleep problems in individuals with MCI, as intrusive negative thoughts may prevent people from sleeping (Fang, et al., 2019). These studies show that the association between depression and self-perceived sleep problems as well as their impact on cognitive decline in individuals with MCI, requires further investigation.

Still, another factor that is commonly associated with sleep problems in older adults is increased stress levels, which can lead to further cognitive decline, particularly in individuals with mild cognitive impairment (MCI). Research has demonstrated a bidirectional relationship between sleep quality and emotional experiences, with increased affective reactivity to stress being a significant predictor of poor sleep quality in older adults. However, after nights of lower sleep quality, older adults report more negative affect, but not higher affective reactivity to stressors (Lücke et al., 2022). In addition, stress-related sleep problems in older adults have been associated with hippocampal atrophy (Wilson et al., 2011), a brain region involved in memory, as well as with the dysregulation of the hypothalamic-pituitary-adrenal (HPA) axis (Martire et al., 2020), both being contributing factors in the deterioration of cognitive functions in people with MCI and potential accelerators in the progression of MCI to dementia (Brown et al., 2014). Investigating this interaction between poor sleep quality and chronic stress is critical for modifying cognitive decline in older adults with MCI.

Finally, quality of sleep is strongly associated with medical health factors and overall health-related quality of life (Leng et al, 2020). Indicative common health-related factors among older adults include cardiovascular-related diseases and metabolic syndrome that appear to also be interwoven with the clinical condition of MCI (Giovanni et al., 2000).

The goal of the current study was to explore the impact of variables related to cognitive functioning, neuropsychiatric symptomatology, and medical health on various sleep problems that individuals with MCI are facing, especially insomnia-related symptoms, by implementing a latent variable approach. More specifically, this was achieved through the development of a partial least square structural equation model (PLS-SEM) that focuses on explaining the variance in the model’s outcome variable and has the capacity to retain its effectiveness even with quite small samples. The selected variables covered important components of psychological and cognitive status, as well as of medical health, such as diabetes mellitus and hypertension. In this context, latent variables were developed to assess psychological and cognitive functioning, as well as to evaluate the outcome measure related to various sleep problems assessed by the Athens Insomnia Scale (AIS). These problems include nighttime awakenings, early final awakening, reduced daytime functioning and well-being, and insufficient total sleep duration. The AIS is considered a suitable tool for this study, as it is specifically designed to assess both night-time sleep problems and daytime dysfunctions resulting from disrupted sleep patterns (Lyrakos et al., 2011; Norton, 2007; Pezirkianidis et al., 2018). This comprehensive assessment of various sleep problems offered by the AIS may prove valuable in deepening the understanding of sleep-related issues in individuals with MCI, a clinical population where insomnia and a broader range of sleep problems are notably prevalent (Hamdy et al., 2018). In addition, the age at onset of MCI was taken under consideration because of the important role of the factor of age on the architectonics of sleep (Casagrande et al., 2022), as well as on the clinical picture of individuals with MCI (Moustaka et al., 2023). To the best of our knowledge, this is the first study that focuses on the sleep problems of individuals with MCI through exploring the unique contribution of a multimodal set of predictors, of which several are treated as latent variables in order to enhance their preciseness, genuineness and overall validity.

## Methods

### Participants

The sample of the present study consisted of 131 patients with MCI (98 females; *M_Diagnosis_age_* = 67.37 ± 8.85; *M_Education___years_* = 12.37 ± 4.03) (Tables 1 & 2), who are consecutive visitors of the Alzheimer’s day clinic - Nestor Psychogeriatric Association. The inclusion criteria were the following: Greek mother language, preserved daily functionality, absence of a psychiatric history or chronic and incurable organic disease. Normal levels of daily functionality were exhibited by participants (see Table 3), while memory or other cognitive complaints had a duration of up to one year before the conduction of the neuropsychological evaluation. Following Petersen and Morris’ criteria, the sample comprised of 90 patients with amnestic multiple-domain MCI, 17 patients with amnestic single-domain MCI, 15 patients with non-amnestic multiple-domain MCI, and 8 patients with non-amnestic single-domain MCI.

**Table 1.** Participants’ Sociodemographic Characteristics (N=131)

| Participants' Characteristics | Number of Participants | Statistics |  |
| --- | --- | --- | --- |
|  | N (%) | Mean (SD) | Range |
| Overall Sample |  |  |  |
| Age | - | 67.37 (8.85) | 44 - 87 |
| Education | - | 12.37 (4.03) | 1 - 20 |
| MMSE | - | 28.06 (1.95) | 20 - 30 |
| IADL | - | 7.90 (1.88) | - |
| Gender |  |  |  |
| Female | 98 (74.8) | - | - |
| Male | 33 (25.2) | - | - |
| Occupational Status |  |  |  |
| Currently Working | 29 (22.5) | - | - |
| Unemployed | 3 (2.3) | - | - |
| Pensioner/Home Economics | 97 (75.2) | - | - |
| Handedness |  |  |  |
| Right-handed | 76 (93.8) | - | - |
| Left-handed | 3 (3.7) | - | - |
| Ambidextrous | 2 (2.5) | - | - |
| Family Status |  |  |  |
| Single | 5 (3.8) | - | - |
| Married | 75 (57.3) | - | - |
| Divorced | 23 (17.6) | - | - |
| Widowed | 28 (21.4) | - | - |

**Table 2.** Participants’ Brief Medical History.

| Medical History | Number of Participants |
| --- | --- |
|  | N (%) |
| Elevated Blood Pressure | 59 (46.8) |
| Diabetes | 23 (18.3) |
| Hypercholesterolemia | 70 (56.0) |
| Stroke | 4 (3.2) |
| Heart Attack | 8 (6.5) |
| Traumatic Brain Injury | 12 (9.6) |
| Smoking | 32 (25.4) |
| Physical Exercise | 19 (15.1) |
| Dementia Medication | 2 (1.6) |
| Psychiatric Medication | 43 (34.4) |
| General Medication | 107 (85.6) |
| Dementia Family History | 47 (38.2) |
| Cardiovascular Disease Family History | 51 (41.5) |

**Table 3.**
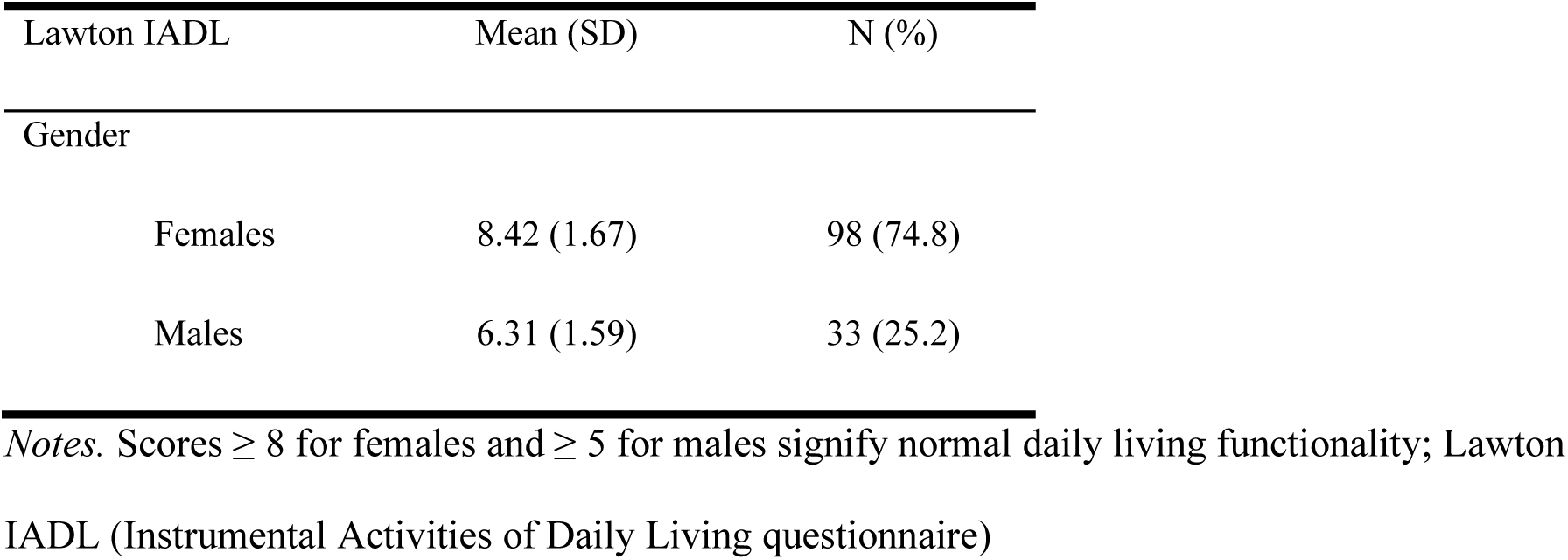
Descriptive Statistics: Levels of Daily Functionality Across Gender Groups.

| Lawton IADL | Mean (SD) | N (%) |
| --- | --- | --- |
| Gender |  |  |
| Females | 8.42 (1.67) | 98 (74.8) |
| Males | 6.31 (1.59) | 33 (25.2) |

*Notes.* Scores $\geq 8$ for females and $\geq 5$ for males signify normal daily living functionality; Lawton IADL (Instrumental Activities of Daily Living questionnaire)

### Materials

The diagnostic procedure of patients consisted of a thorough evaluation, which included the following well-established and valid instruments.

#### Athens Insomnia Scale (AIS-8)

This 8-item scale examines insomnia severity based on the International Classification of Diseases (ICD-10; Soldatos et al., 2000). The following eight domains are assessed: sleep onset, time, and quality, night- and early-morning waking, frequency and duration of complaints, distress experienced due to insomnia, and interference with daily functioning. Scores may range from 0 (no problem) to 3 (more acute sleep difficulties), with an overall cutoff score of 6. Good levels of reliability have been revealed for AIS-8 (Cronbach’s alpha of 0.806).

#### Depression Anxiety Stress Scale (DASS-21)

This scale assesses a three-dimensional level of psychological burden (depression, anxiety, and stress) (Lyrakos et al., 2011; Norton, 2007; Pezirkianidis et al., 2018). The depression dimension evaluates lack of interest/participation, despair, dysphoria, self- and life-depreciation, and anhedonia. The anxiety dimension examines arousal of the subjective experience of anxiety, state anxiety, as well as the autonomous nervous system and its musculoskeletal effect. The stress dimension evaluates irritability/hyperreactivity, temperament, hyperarousal, restlessness, and impatience. Each dimension contains seven items, with scores ranging from 0 (none) to 3 (very much). Each DASS dimension/subscale has been presented with good levels of reliability (Cronbach’s alpha: Depression: 0.929, Anxiety: 0.945, Stress: 0.923) (Ali et al., 2022).

#### Lawton–Brody Instrumental Activities of Daily Living (IADL)

This 8-item scale examines patients’ level of autonomy in daily functioning (Latwon & Brody, 1969). A performance-adequacy index related to the following eight daily functions is computed: ability to make/receive telephone calls, shopping, preparation of meals, housekeeping, laundry, mode of transportation, own-medication responsibility, and management of finances. This scale has good levels of reliability (Cronbach’s alpha: 0.809).

#### Mini Mental State Examination (MMSE)

This scale is a globally used measure of general cognitive status (Folstein et al., 1975; Fountoulakis et al., 2000), that examines attention, language, time and space orientation, memory, and visuospatial skills. Scores may range from 0 (very severe cognitive impairment) to 30 (no cognitive impairment). Good levels of reliability have been published for the MMSE (Cronbach’s alpha: 0.780) (Kabatova et al., 2016).

#### REY Auditory Verbal Learning Test (RAVLT)

This scale assesses individuals’ capacity to encode, consolidate, store, and retrieve verbal information. It consists of five presentation trials of a 15-word list of nouns (list A), followed by a single presentation of a 15- noun interference list (list B), and two post-interference recall trials (one immediate and one delayed). A final recognition trial is presented, consisting of 50 words from both lists A and B, including 20 distractor words, that are semantically or phonemically similar to the latter lists’ words (Lezak et al., 2004; Messinis et al., 2007; Rey, 1964; Schmidt, 1996). High reliability levels have been shown for this scale (Cronbach’s alpha: 0.917).

#### Trail Making Test A & B (TMT A & B)

This test examines executive functioning, visual search and processing speed, and mental flexibility, consisting of parts A and B (Reitan, 1979). In part A, the individual needs to link a series of numbers (1–13) in ascending order by drawing consecutive lines within 180 s. In part B – a measure of executive control – the individual is required to connect a series of numbers (1–13) and letters (A-L/A-Μ in the Greek version) in an ascending, alternating pattern (e.g., 1-A-2-B-3-C-4-D, etc.) within 300 s. Both tasks must be completed as quickly as possible, without lifting the pencil or pen from the paper. The final score may be affected by possible errors made during the tasks.

#### Verbal Fluency Test (VFT)

The VFT evaluates functions related to the brain’s frontal and temporal lobes, consisting of the following two components: phonemic fluency (PF) and semantic fluency (SF). Analogously to the Benton and Hamsher’s (1976) PF-related use of English letters “F”, “A”, and “S”, the examinee is required to elicit words that begin with the Greek letters “Χ”, “Σ”, “Α”, within three minutes (one minute per letter) (Kosmidis et al., 2004). Likewise, in SF, the examinee is asked to evoke words that fit in one of the three categories (animals, fruits, and objects), within three minutes (one minute per category) (Kosmidis et al., 2016; Strauss et al., 2006). Both components of this scale have exhibited very good levels of reliability (Cronbach’s alpha: PF: 0.873, SF: 0.767).

### Procedure

The present study included participants who were consecutive visitors of the Outpatient Memory Clinic of Nestor Alzheimer’s Centre. The inclusion criteria were the following: complaints regarding cognitive dysfunction, Greek mother language, absence of psychiatric history, and a valid MCI diagnosis based on Petersen’s criteria (Petersen et al., 2001). A thorough neuropsychological and medical assessment was completed by all participants, after providing a written consent. A team of professional psychologists, neuropsychologists, and psychiatrists conducted 120-minute evaluations, that included socio-demographic questions, and a brief medical history, as well as, administering the following questionnaires: Lawton– Brody IADL, MMSE, RAVLT, VFT (both PF and SF), Trail Making Test (TMT, both parts A and B), and DASS-21. It must be noted that the Greek standardized version of each scale was utilized for data collection.

### Ethical Considerations

Both institutions involved in this research, including (a) the Psychology Ethics Research Committee, operating under the auspices of the Institutional Review Board at the American College of Greece, and (b) the Ethics Committee of the Alzheimer’s Outpatient Day Center, “Nestor” Greek Psychogeriatric Association, approved the present study. Through the informed consent form, all individuals were notified about their right to withdraw at any time, the confidentiality of the procedure, and that the use of their background information would be only for research purposes. Participants were further informed about the nature and the engagement duration of the study, as well as about the type of information to-be-asked during the data collection process. Participation was voluntary, and no compensation was offered.

### Statistical Analyses - Model Estimation

All latent variables of the model were operationalized as reflective. More specifically, the latent variable “psychological burden” included as indicators the total scores of depressive symptoms, anxiety, and stress, as they are assessed by the DASS. The latent variable “cognitive status” included as indicators the total MMSE score, the total phonemic fluency and semantic fluency scores, the completion time of the TMT-A and the 5^th^ trial of the Rey-AVLT. Finally, the latent variable “sleep problems” included as indicators the following symptoms of the Athens Insomnia Scale: a) problems with awakening during night, b) earlier final awakening, c) decreased functioning capacity during the day, d) decreased well-being during the day, and e) insufficient total sleep duration. For all latent variables the indicators that were selected should produce a composite reliability index (rho_c) above the level of 0.7 for sufficient levels of internal consistency to be present and the Average Variance Extracted (AVE) should be above the threshold of 0.5 for convergent validity to be sufficiently met. To estimate our model, we applied a Partial Least Squares (PLS) approach to structural equation modeling using SmartPLS 4 (Ringle et al., 2024). Bootstrapping procedures were run with 10000 subsamples.

## Results

For the latent variable “Psychological Burden” the outer loadings of the indicators are above the level of 0.7 and the composite reliability index (rho_c) is equal to 0.895 (well above the threshold of 0.7) indicating high internal consistency. In addition, the Average Variance Extracted (AVE) is above the threshold of 0.5 (AVE=0.741) suggesting that convergent validity is met. For the latent variable “Cognitive Status” the outer loadings of the indicators are not reaching in all cases the optimal level of 0.7. Nonetheless, all values are above the level of 0.58 (range: 0.581-0.849) and the composite reliability index is above the level of 0.7 (0.707). In addition, AVE is 0.522, suggesting that convergent validity is met. For the latent variable “sleep problems” the outer loadings of the indicators are not reaching in all cases the optimal level of 0.7. Nonetheless, all values are above the level of 0.553 (range: 0.553-0.817) and the composite reliability index is equal to 0.839 (well above the threshold of 0.7). In addition, AVE is above the threshold of 0.5 (AVE=0.514), suggesting that for this latent variable convergent validity is also met. Also, discriminant validity is met since the Heterotrait-Monotrait (HTMT) Ratios of all constructs of the model are well below the threshold of 0.9 (the maximum HTMT ratio that was observed was equal to the value 0.661 in the upper bound [95%]).

Problems of multicollinearity are not identified regarding the structural model, since all VIF values are below the level of 3 (maximum VIF value in the model is equal to 2.061). Also, all indicators are reaching the level of statistical significance (sig. value < 0.05) and their outer loading is above the level of 0.5. Regarding the predictive power of the model, all PLS Root Mean Square Error (PLS_RMSE) values are lower from the respective Linear-Regression Model Root Mean Square Error (LM_RMSE) values and, therefore, the model has high predictive power. In addition, The Stone-Geisser Q^2^ values obtained through the blindfolding procedures were larger than zero, supporting the predictive relevance of the model (Hair Jr. et al., 2017). Finally, a good model fit appears to be present because the standardized root means square residual (SRMR) value was below the threshold of 0.08 (SRMR=0.064) (Hair Jr. et al., 2017).

Regarding the main findings, as seen in Figure 1, the effect size of psychological burden (f^2^=0.097) is clearly larger as compared to the effect size that was observed in the case of cognitive status (f^2^=0.028). In this direction, significant results were observed for the following predictors: psychological burden (*b*=0.296, *p*<.001), cognitive status (*b*=-0.183, *p*=0.011), and the age of onset of MCI (*b*=-0.152, *p*=0.012). More specifically, for the latent variable “psychological burden” that reflects symptoms related to depression, anxiety and stress a positive association was observed with the severity of the sleep problems. In the case of the latent variable “cognitive status” a negative association was observed with “sleep problems”. Finally, a negative association was detected between the age of onset of MCI and the density of sleep problems.

**Figure 1.**
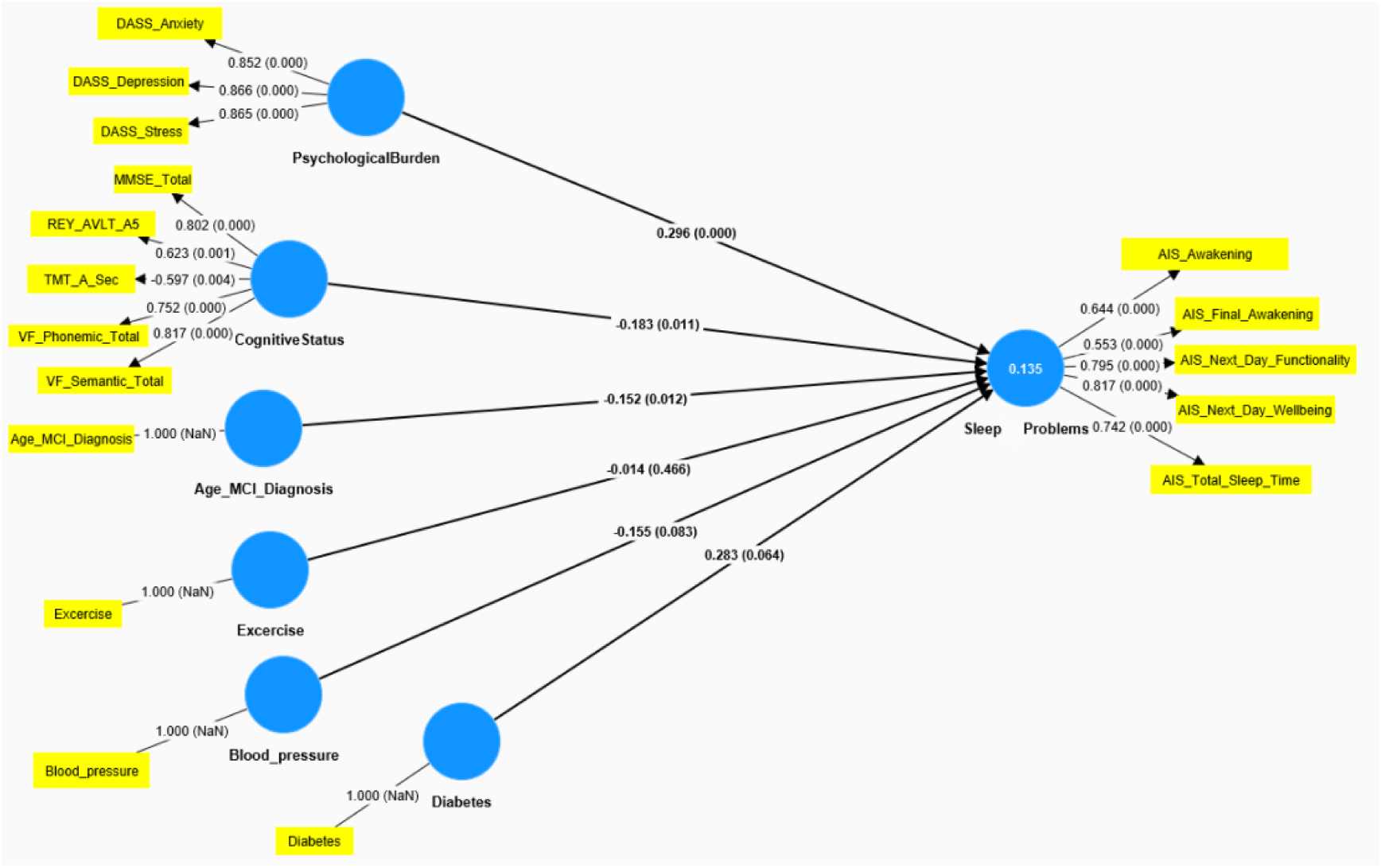

## Discussion

The SEM approach, which was implemented for the needs of the current study, made it feasible to explore the impact of a broad array of first-level and latent variables on the development of sleep problems in individuals with MCI. By utilizing a latent variable approach our goal was to assess the relevant constructs in a more accurate and holistic way, as well as to produce more reliable and valid measurements by reducing the amount of measurement error that is observed in the case of the first-level variables. In this direction, the corresponding latent variable that focused on the assessment of various sleep problems and served as the outcome measure included as indicators the following symptoms that were extracted from the AIS: a) problems with awakening during the night, b) earlier final awakening, c) decreased functioning capacity during the day, d) decreased well-being during the day, and e) insufficient total sleep duration. The obtained results supported the effectiveness of the constructed model since the overall model had a good fit and a high predictive power. Among the assessed predictors, the latent variable “psychological burden”, which included depression, anxiety, and stress symptoms, appeared to clearly have the most important role in the development of sleep problems in the specific clinical group. The second most important predictor of the model was the latent variable “cognitive status” which covered various cognitive domains, namely general cognitive status, episodic memory, verbal fluency, and psychomotor speed. According to the negative association that was observed in this case, it appears that individuals with MCI, who manifest an accelerated cognitive burden, may have an increased risk of developing sleep problems as well. Finally, a negative association was detected between the age of onset of MCI and the severity of sleep problems, thus indicating a greater tendency for sleeping difficulties in individuals who develop MCI at a younger age. Finally, non-significant results were observed in the case of the following predictors: physical exercise, the presence of diabetes mellitus, and the presence of high blood pressure.

The results of our study could provide us with some meaningful insights into psychological burden, cognitive status, and sleep disturbance progression in MCI. Psychological burden emerged as the strongest predictor of sleep problems, followed by cognitive functioning. These findings are consistent with previous evidence that highlights how neuropsychiatric symptoms often dominate the clinical presentation of aging adults with cognitive impairment and have a significant impact on sleep quality in this group. Previous studies have shown that depression and anxiety are widespread among older adults with MCI (Gallagher et al., 2017; Ismail et al., 2017; Martin & Velayudhan, 2020; Chen et al, 2018; Martin & Velayudhan, 2020), and are often associated with sleep problems, contributing to poor outcomes, including elevating the risk of cognitive decline (Norton et al., 2014; Xu, et al., 2019). These studies primarily evaluated individual neuropsychiatric factors, focusing on either depression or anxiety and found a strong association with sleep quality in MCI patients (Seidel et al; 2014; Song et al, 2020). Similarly, stress has been shown to negatively affect sleep quality and further contribute to cognitive decline (Lücke et al., 2022). However, few studies have specifically examined this relationship in MCI patients (Brown et al., 2014), with most research focusing on the general population of older adults (Wilson et al., 2011). Including stress as a component in our study is particularly beneficial, given the limited research on its role in sleep problems among individuals with MCI. Our results build on these observations by providing a more refined understanding, of where psychological burden, when assessed as a composite variable, considerably predicts sleep problems in individuals with MCI.

The negative association between cognitive status and sleep problems provides additional evidence for the hypothesis that declining cognitive status is a significant risk factor for the exacerbation of sleep problems. Our finding is also in line with Xu et al. (2020), who reported that sleep problems (especially insomnia) are associated with a higher risk of MCI, and later, of dementia. The inverse association found in our study suggests that deterioration in some cognitive functions, especially in episodic memory, verbal fluency, and psychomotor speed, increases the risk for sleep problems. The interwoven nature of sleep problems and cognitive symptoms might facilitate a process of reciprocal influence, where the two clinical conditions mutually trigger each other, resulting in an exacerbation of the pathological process underlying MCI, thus leading more quickly to dementia. Along this vein, the findings of a previous study have shown that sleep problems mirror the evolution of cognitive impairment in individuals with MCI and Alzheimer’s disease (Casagrande et al., 2022).

While both latent variables, “psychological burden”, and “cognitive status”, predict sleep problems, our findings indicate that “psychological burden” is a stronger predictor. This may be attributed to the widespread impact of psychological symptoms, such as depression, anxiety, and stress, which tend to be more prominent and exert a more direct influence on sleep quality than cognitive decline alone (Wilson et al., 2011). The stronger predictive value of psychological burden shows the need to consider emotional and psychological factors when managing sleep problems and to incorporate them into the overall treatment plans for individuals with MCI.

The nonsignificant findings regarding sleep problems in relation to physical exercise, diabetes mellitus, and high blood pressure are important, given the established literature on these factors too. Whereas hypertension, obesity, and lack of physical exercise are indeed known to be risk factors that are significantly associated with cognitive decline (e.g., Norton et al., 2014), such associations were not found in the current study to produce a significant impact on the levels of sleep problems of individuals with MCI. Hence, despite the role of these factors on the observed cognitive burden of individuals with MCI, their impact on the sleep problems does not appear to be equally important because of the nonsignificant results that were obtained in the current study. Nonetheless, implementing an alternative methodological strategy that enables a detailed evaluation of various health-related domains could be a reasonable target for future research. This approach would facilitate the development of a latent variable structure for this set of predictors, providing clearer and more solid conclusions about the impact of physical health on the sleep problems of this specific clinical group.

Interestingly, we found that older age at onset of MCI was independently associated with less severe sleep problems, though there may be multifold explanations for this. It could be argued that those who develop MCI at an older age have a less aggressive underlying pathology (Ho et al., 2002; Jacobs et al., 1994; van der Vlies et al., 2009; Wattmo & Wallin, 2017). Ye and colleagues (2012) showed significant differences in the neuropsychological performance and disease progression between early-onset and late-onset MCI. Individuals with early-onset aMCI demonstrated a broader range of cognitive impairments and a higher likelihood of converting to AD, compared to late-onset. Alternatively, one may argue that older adults who develop MCI are less aware of and less likely to objectively report sleep problems, thus leading to lower measured severity. From a clinical standpoint, this suggests that younger patients with MCI should be targeted for earlier intervention to manage sleep problems and prevent disease progression.

Although the psychological burden-related measures were based on in-depth evaluations, that was not the case for variables focusing on medical symptoms and physical exercise, which were based on single-question assessments. Thus, we were not able to study specific physical-health-related measures as a latent variable – a step that may provide further information about the overall health status of individuals and its connections to their levels of psychological burden and sleep problems. Furthermore, the use of additional assessment scales of sleep problems may also enhance the strength of the latent-variable model through the increased number of sleep-related sub-variables measured. Targeting the use of a broader spectrum of evaluation tools of psychological, physical, and sleep-related measures, could possibly help future researchers to gain more insights from latent models about the level and the way that each factor may impact sleep in individuals with MCI.

Overall, by recognizing the complexity of sleep problems in MCI, we were able to use a latent variable model in order to explore the unique contribution of a broad spectrum of predictors. Along this vein, the current study provides further evidence of the significant impact of neuropsychiatric symptoms, such as depression, anxiety, and stress, on the sleep problems experienced by individuals with MCI. Additionally, it supports the distinct role that cognitive burden plays in contributing to the sleep difficulties within this clinical group. This combined pattern of results highlights the significance of integrative assessments and interventions that effectively target the psychological wellbeing and cognitive status of MCI patients to improve their sleep quality and potentially slow down the progression of their cognitive decline.

## Data Availability

All data produced in the present study are available upon reasonable request to the authors.

## Author Contributions

I.N.B. made substantial contributions to the conception and design of the study, the analysis and interpretation of data, drafting and revising it critically for important intellectual content, and gave final approval of this version to be published. He agrees to be held accountable for all aspects of this study and ensure that questions related to the accuracy or integrity of any part of this study are appropriately investigated and resolved. K.M. made substantial contributions to the collection of data and the conception of the study, drafting and revising it critically for important intellectual content, and gave final approval of this version to be published. She agrees to be held accountable for all aspects of the work and ensure that questions related to the accuracy or integrity of any part of this study are appropriately investigated and resolved. C.N. made significant contributions to the present study through its conception and design, drafting and revising it critically for important intellectual content, and gave final approval of this version to be published. She agrees to be held accountable for all aspects of this study and ensure that questions related to the accuracy or integrity of any part of this study are appropriately investigated and resolved. All authors have read and agreed to the published version of the manuscript.

## Institutional Review Board Statement

The study was conducted in accordance with the Declaration of Helsinki and was approved by: (a) the Ethics Committee of the Alzheimer’s Outpatient Day Center, “Nestor” Greek Psychogeriatric Association, and (b) the Psychology Ethics Research Committee, operating under the auspices of the Institutional Review Board of The American College of Greece.

## Informed Consent Statement

Individual informed consent forms were signed by all participants of the present study, allowing the researchers to carry out statistical analyses of the anonymized clinical data.

## Data Availability Statement

For data availability and access, please contact.

